# Sex and Obesity Stratified Asthma GWAS in African and European Ancestry Populations

**DOI:** 10.64898/2026.07.05.26357321

**Authors:** Hui-Qi Qu, Michael March, Frank Mentch, Haijun Qiu, John J Connolly, Joseph T Glessner, Hakon Hakonarson

**Affiliations:** The Center for Applied Genomics, Children’s Hospital of Philadelphia, Philadelphia, Pennsylvania, 19104, USA; Department of Pediatrics, The Perelman School of Medicine, University of Pennsylvania, Philadelphia, Pennsylvania, 19104, USA; Division of Human Genetics, Children’s Hospital of Philadelphia, Philadelphia, Pennsylvania, 19104, USA; Division of Pulmonary Medicine Children’s Hospital of Philadelphia, Philadelphia, Pennsylvania, 19104, USA; Faculty of Medicine, University of Iceland, 101 Reykjavik, Iceland

**Keywords:** asthma, genome-wide association study, obesity, sex, stratified analysis, African ancestry, European ancestry

## Abstract

**Background:** Biologically distinct asthma subgroups may obscure genetic effects when analyzed as a single phenotype. We examined whether asthma susceptibility signals are shared, heterogeneous, or stratum-specific across ancestry, obesity status, and sex.

**Methods:** We performed ancestry-specific GWAS meta-analyses in African ancestry participants (9,965 asthma cases; 37,391 controls) and European ancestry participants (6,074 cases; 116,255 controls), followed by obesity- and sex-stratified analyses. Analyses used imputed dosages and fixed-effect meta-analysis within ancestry.

**Results:** Stratification detected asthma association signals that were less apparent in the combined phenotype. Shared cross-ancestry loci implicated epithelial antiviral susceptibility and immune regulation, represented by signals near *CDHR3* and *FOXO1*. An ancestry-heterogeneous signal at the 17q21 locus, harboring *ORMDL3*/*GSDMB,* supported population-dependent effects at an epithelial inflammatory locus. Obesity stratification mapped the genome-wide significant burden to asthma without obesity. Sex stratification detected genome-wide significant signals in AFR females with asthma and obesity and in both sex strata with asthma without obesity, with the strongest signal burden in EU females without obesity.

**Conclusions:** Asthma genetic architecture differed by ancestry, obesity status, and sex. Stratified analyses identified group-specific susceptibility related to epithelial and immune regulation, airway inflammation, remodeling, and neural signaling, supporting precision approaches to asthma.

## Introduction

Asthma is a heterogeneous airway disease in which genetic susceptibility acts within epithelial, immune, environmental, and physiological contexts. GWAS have identified asthma susceptibility loci related to epithelial antiviral defense, type 2 inflammation, immune-cell regulation, and airway remodeling (1, 2). Some asthma signals are shared across populations, whereas others differ with allele frequency and linkage disequilibrium (LD) structure, or population-specific immune and airway biology (3).

In addition to ancestry, obesity and sex are key biological states that modify asthma susceptibility. Obesity reduces lung volume, promotes airway narrowing, and predisposes to low-grade systemic inflammation. Obesity-related adipokines and comorbid metabolic conditions can also worsen asthma control (4). These processes alter the inflammatory state in which asthma susceptibility alleles act. Asthma prevalence is higher in boys during childhood and higher in women after puberty, with sex hormones contributing to differences in asthma incidence, severity, and inflammatory response (5). Estrogen and progesterone can promote type 2 inflammation, eosinophilic activity, and airway hyperresponsiveness, whereas androgens tend to suppress type 2 immune responses (6, 7). Sex hormones also affect airway epithelial signaling and remodeling, creating sex-specific inflammatory contexts in which asthma risk alleles may act (8).

Most asthma GWAS analyses do not separate biologically distinct subgroups, although obesity status and sex are often modeled as covariates. This approach may obscure susceptibility signals that differ across these biological states. In this study, we performed ancestry-specific asthma GWAS meta-analyses in African (AFR) and European (EU) ancestry participants, followed by obesity- and sex-stratified analyses. We evaluated shared variants, ancestry-, obesity-, and sex-based heterogeneous effects, and genome-wide significant (GWS) loci across the stratified asthma dataset at the Center for Applied Genomics at the Children’s Hospital of Philadelphia.

## Methods

### Study design

This study was conducted separately in AFR and EU subsets. Genotype data were analyzed as imputed dosages rather than best-guess genotype calls. The AFR dataset included multiple genotyping array or imputation batches, with 9,965 asthma cases (5,437 males; 4,528 females) and 37,391 controls (16,456 males; 20,935 females). The EU dataset included comparable genotyping array or imputation batches, with 6,074 asthma cases (3,498 males; 2,576 females) and 116,255 controls (60,150 males; 56,105 females). Each batch contained data from a single genotyping array, while some arrays were split across multiple imputation batches.

Genome-wide association analyses were first performed for all asthma cases versus controls. Obesity-stratified analyses were then conducted for asthma with obesity and asthma without obesity within each ancestry group. In AFR, asthma with obesity included 2,483 cases (1,211 males; 1,272 females) and 2,618 controls (1,010 males; 1,608 females), whereas asthma without obesity included 7,482 cases (4,226 males; 3,256 females) and 34,773 controls (15,446 males; 19,327 females). In EU, asthma with obesity included 657 cases (359 males; 298 females) and 1,066 controls (515 males; 551 females), whereas asthma without obesity included 5,417 cases (3,139 males; 2,278 females) and 115,189 controls (59,635 males; 55,554 females). Sample details by ancestry, phenotype, sex, and genotyping or imputation batch are provided in Supplementary Tables 1 and 2.

### Genotyping, quality control, and imputation

Samples were genotyped on Illumina arrays, including the Global Screening Array, HumanHap, Omni25, and OmniExpress/OmniExpressExome-derived merged datasets (Illumina, CA, USA). Genotype data were processed in batch-specific datasets defined by array and imputation subset. Alleles were aligned to the reference panel before imputation. Genome-wide imputation was performed on GRCh38 using the TOPMed Imputation Server, version R2 (9).

### Genetic association analysis

Association analyses were performed separately in AFR and EU ancestry groups using PLINK v2.0 (10) and imputed allele dosages. Principal components (PCs) were estimated within ancestry from quality-controlled genotype data. Analyses were conducted for overall asthma and for obesity- and sex-stratified asthma phenotypes. Within each ancestry group and genotyping or imputation batch, association testing used logistic regression in PLINK v2.0, adjusting for age, sex, and the first six PCs. Sex was omitted from sex-stratified models. Continuous covariates were variance-standardized before model fitting. Binary phenotypes were tested using logistic regression with Firth fallback for variants requiring bias-reduced estimation.

### Imputation-batch-level meta-analysis

Batch-level summary statistics were combined across batches within each ancestry group using inverse-variance weighted fixed-effects meta-analysis. Batches were analyzed separately to retain batch-specific imputation uncertainty in dosage-based association statistics even when the underlying genotyping array was the same (11). Meta-analyses were performed for each asthma phenotype in AFR and EU.

## Results

### Asthma GWAS

Ancestry-specific fixed-effect meta-analyses identified GWS asthma loci in both AFR and EU participants, with more GWS variants and lead loci in EU. For cross-ancestry sharing, one AFR GWS lead locus showed nominal same-direction evidence in EU, and three EU GWS lead loci showed nominal same-direction evidence in AFR after allele harmonization (Table 1; Supplementary Table 3). Among variants with significant AFR-EU heterogeneity, 302 were suggestive in at least one ancestry, including 49 variants suggestive in AFR and 253 suggestive in EU (Supplementary Table 4). Of the EU suggestive heterogeneous variants, 79 also reached GWS in EU. No significantly heterogeneous variants were suggestive or GWS in both ancestries.

**Figure 1.**
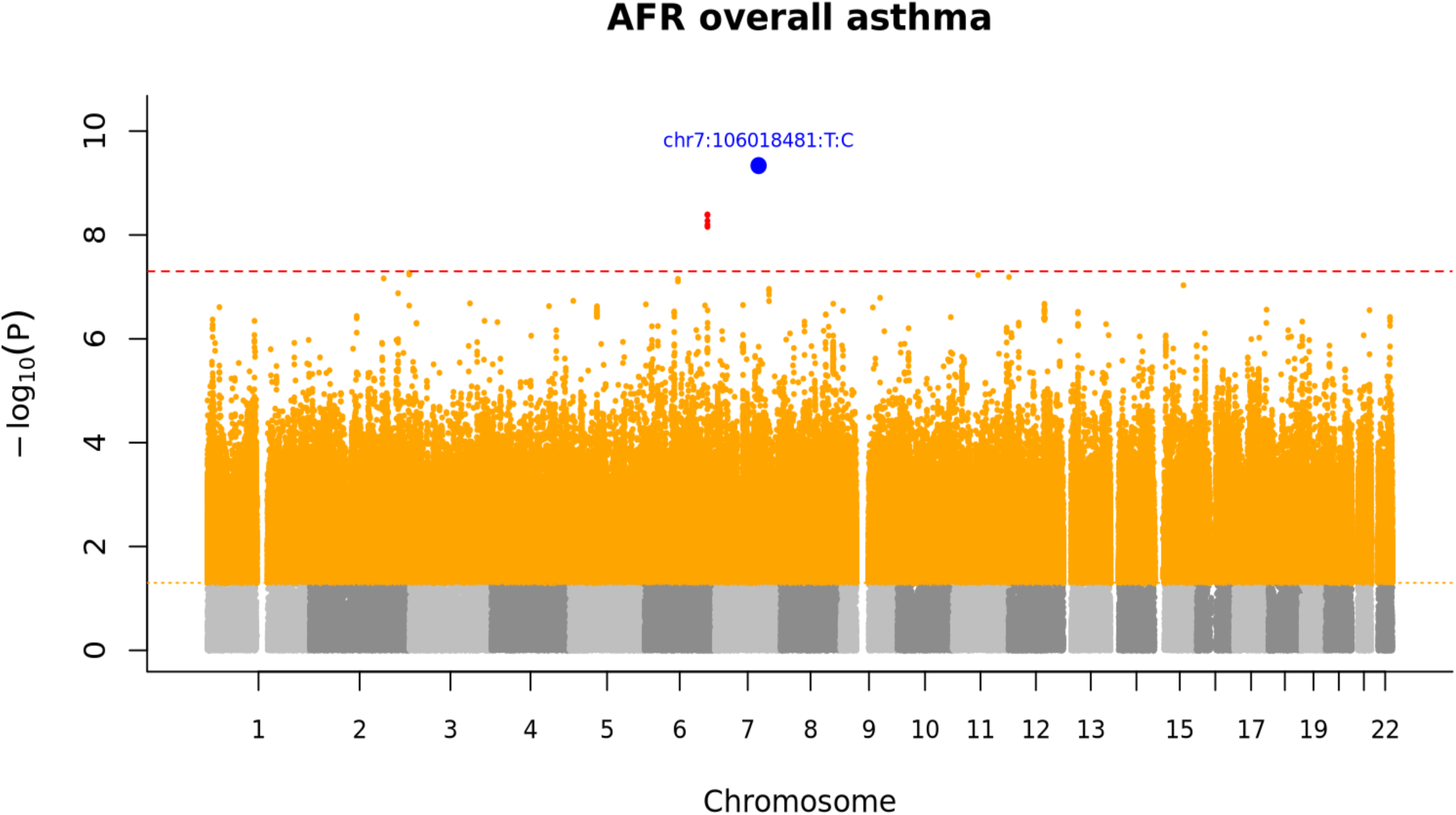
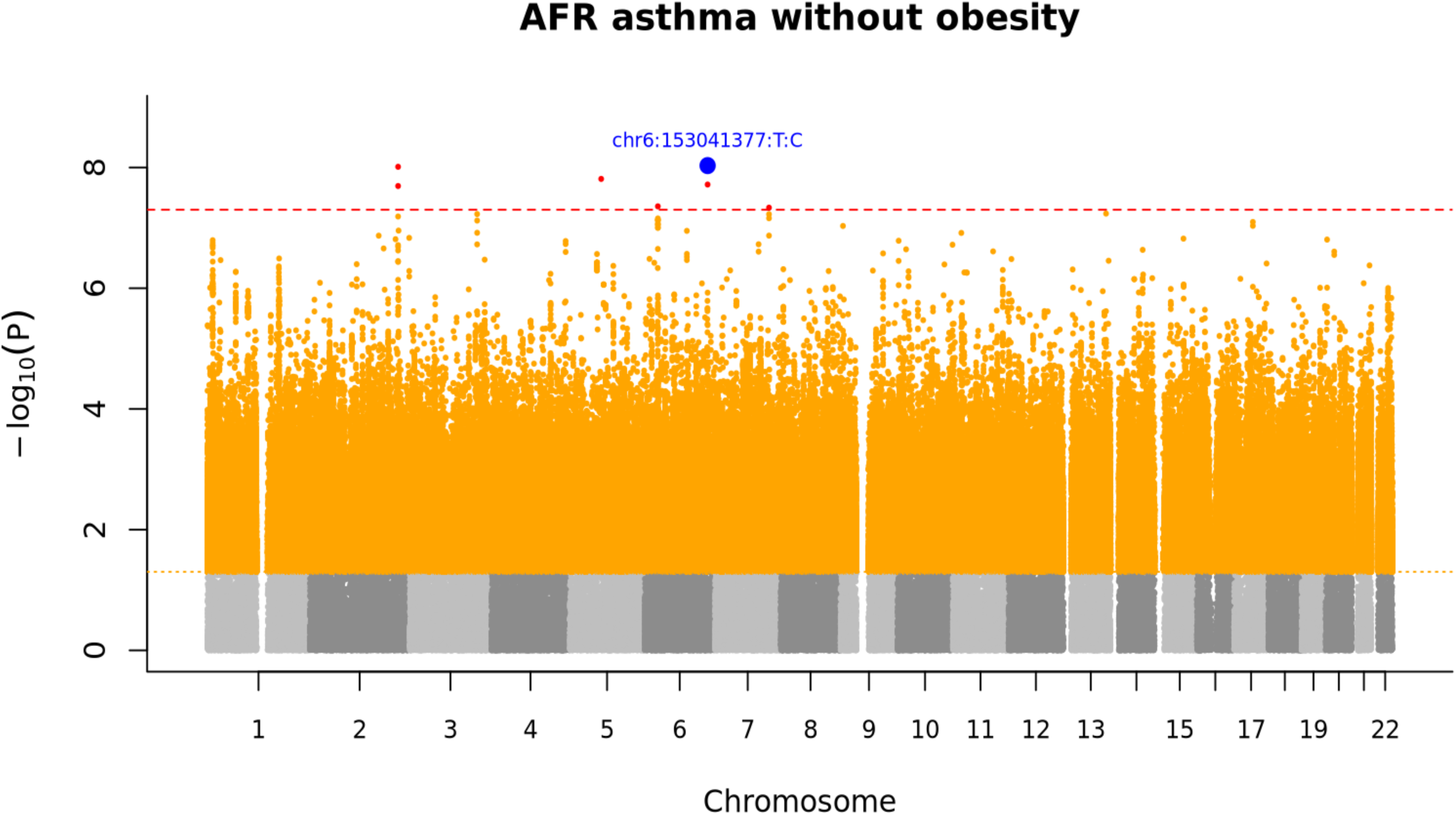
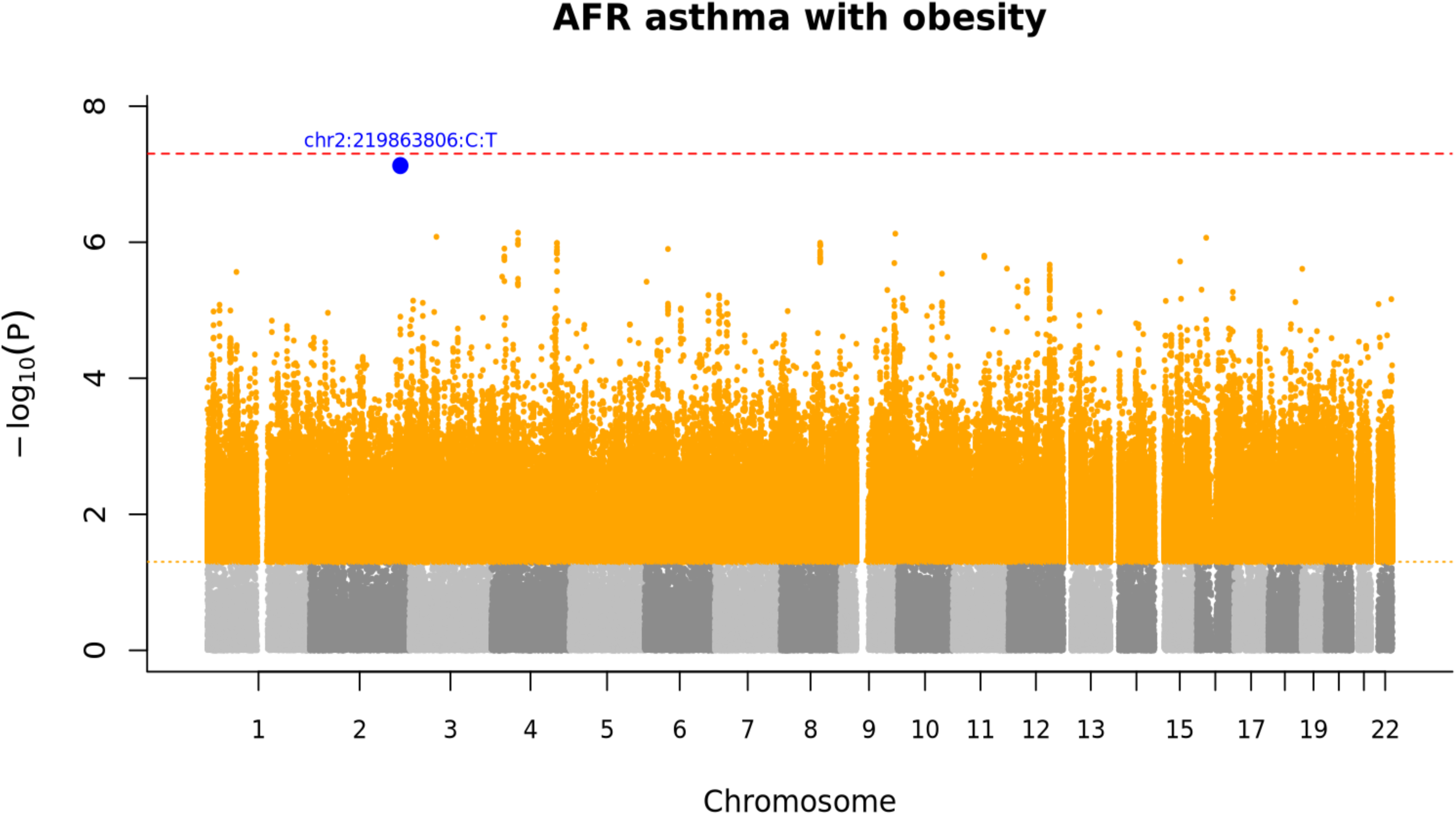
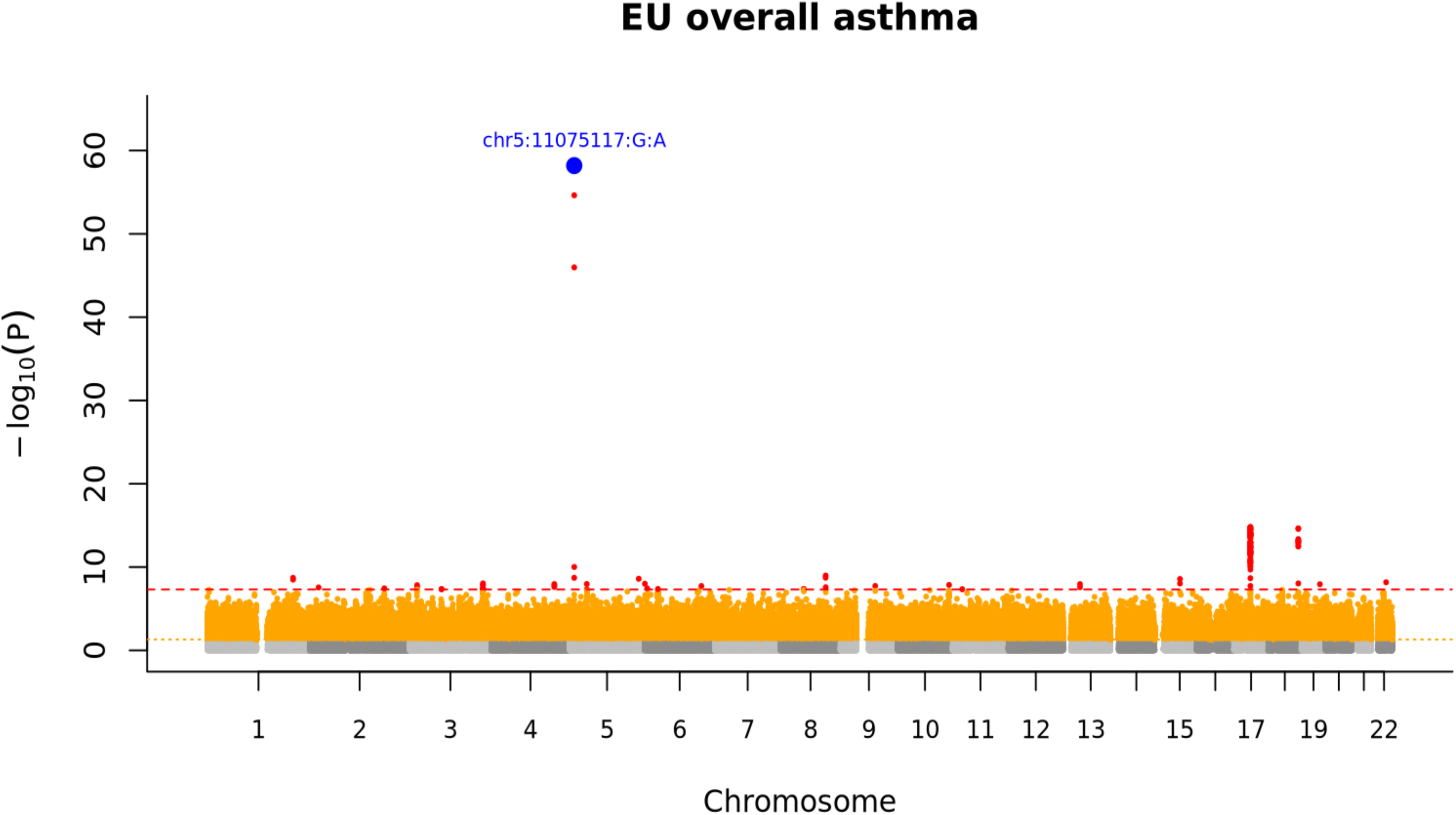
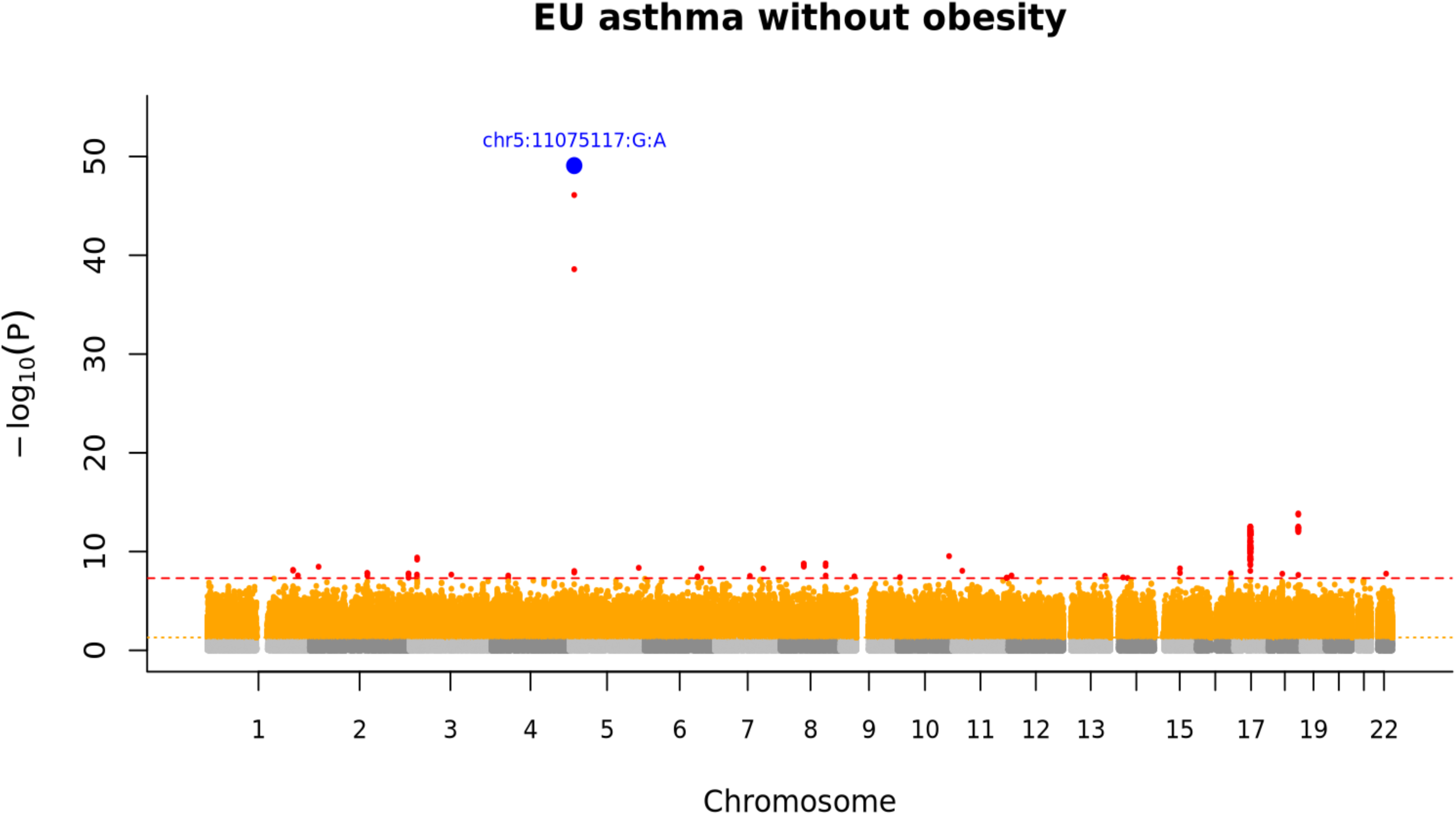
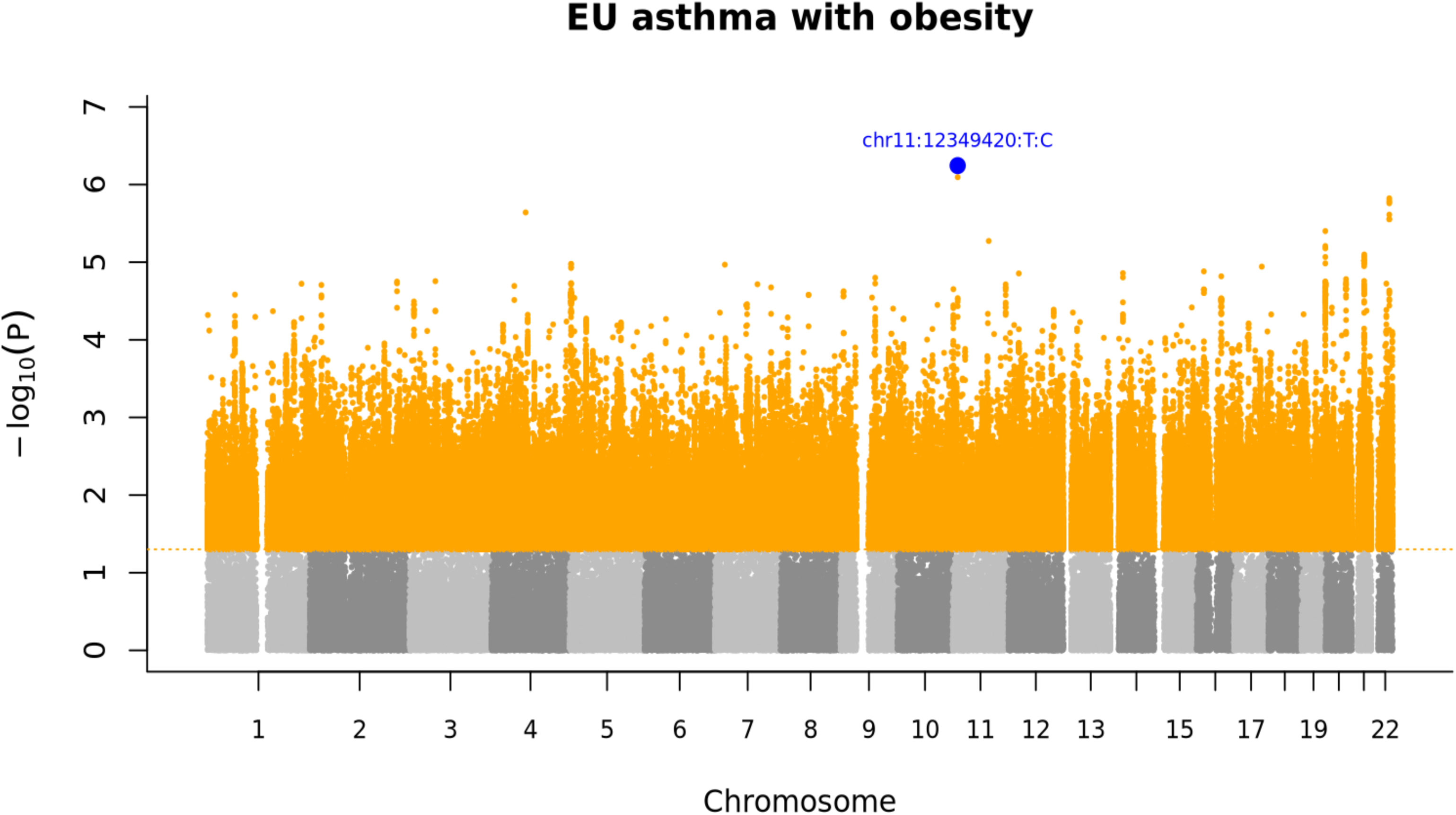
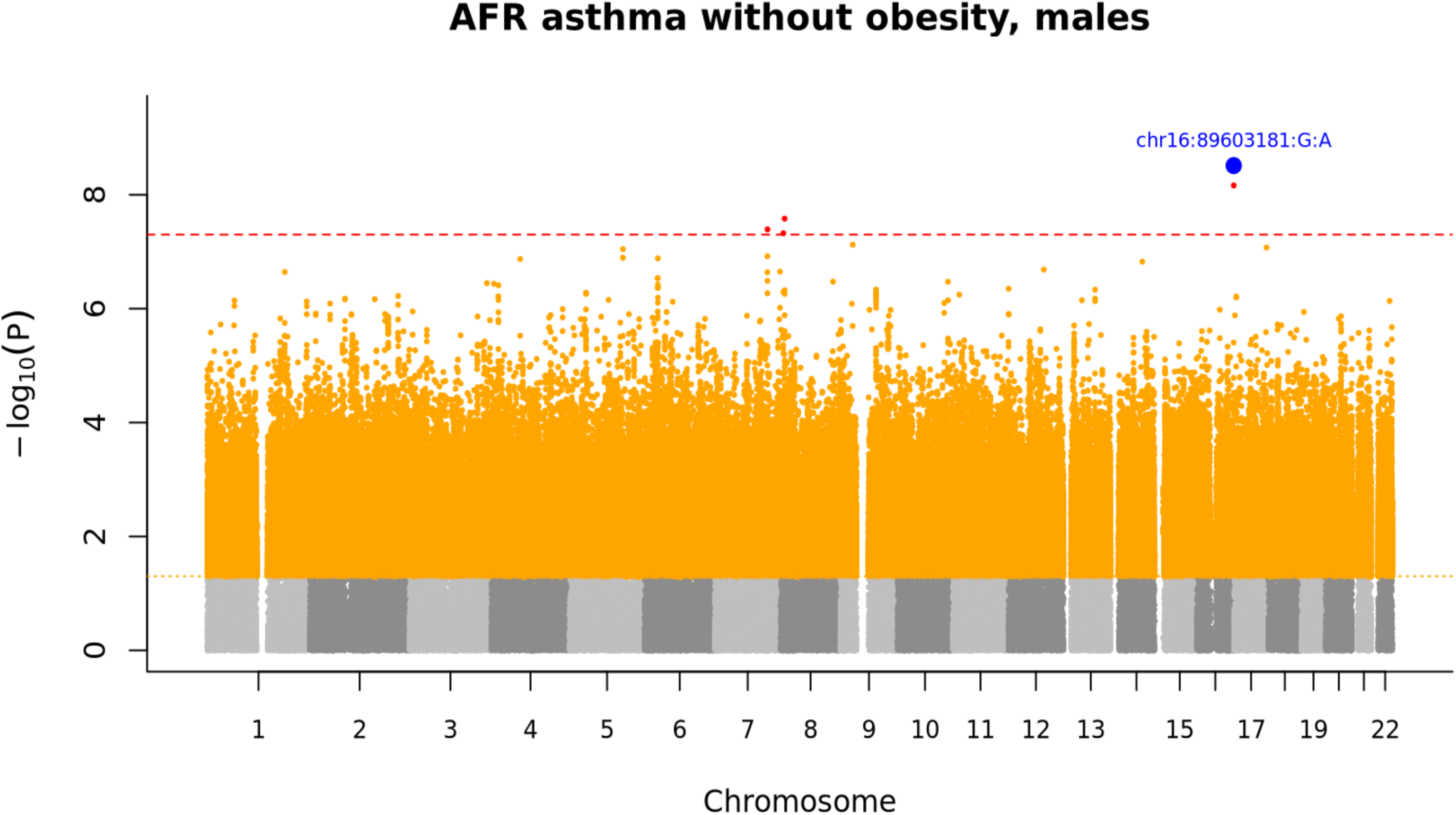
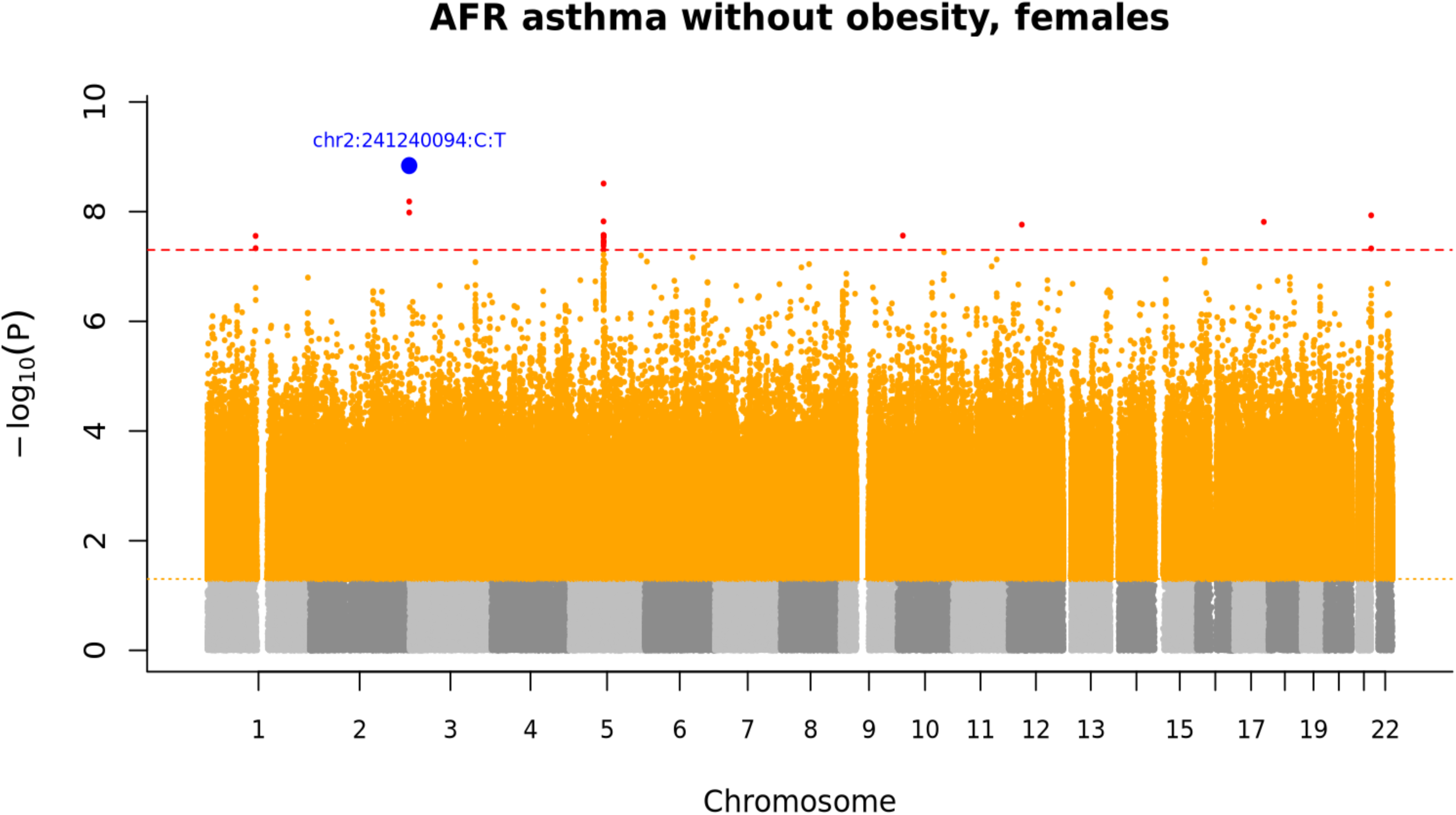
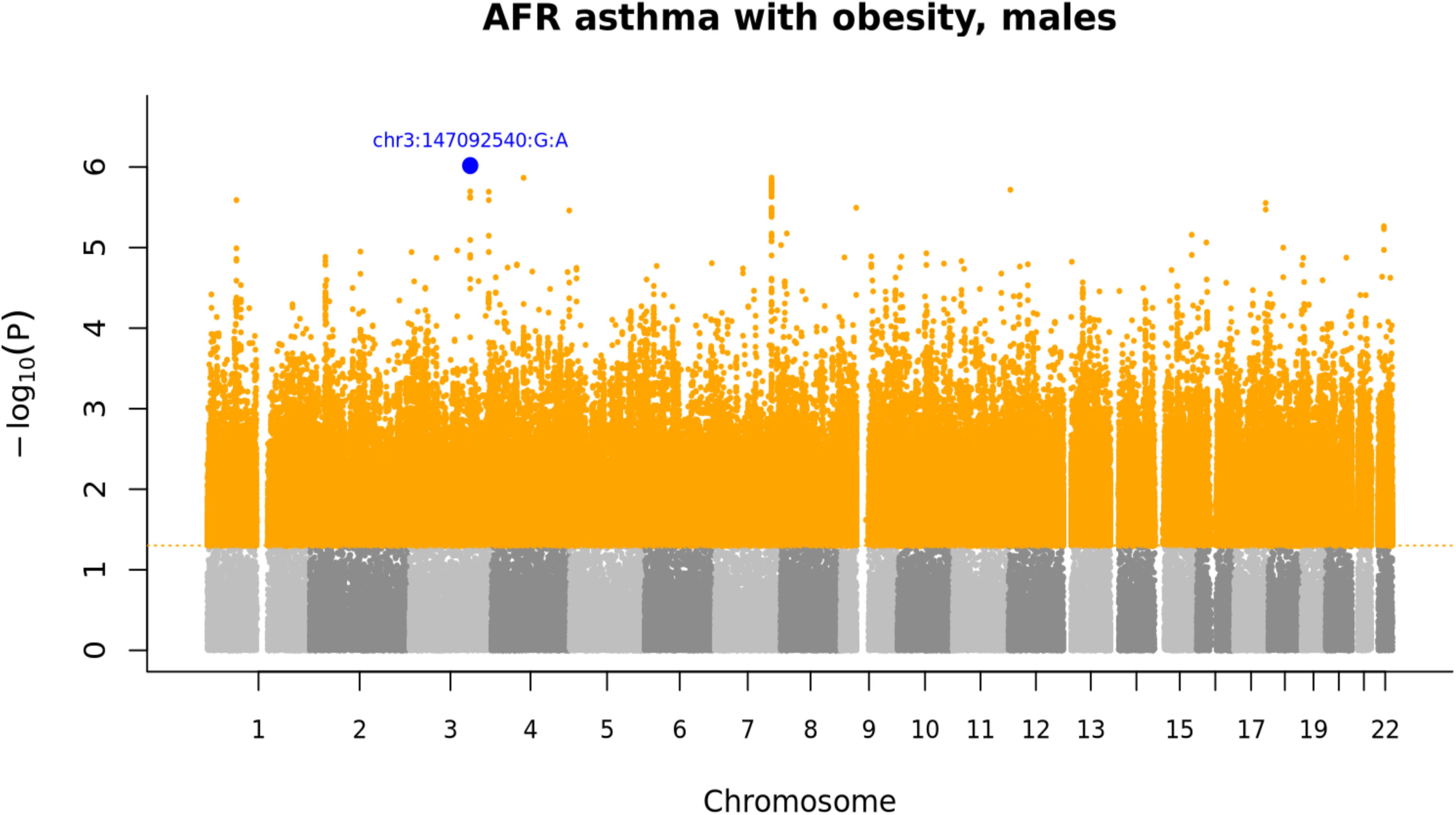
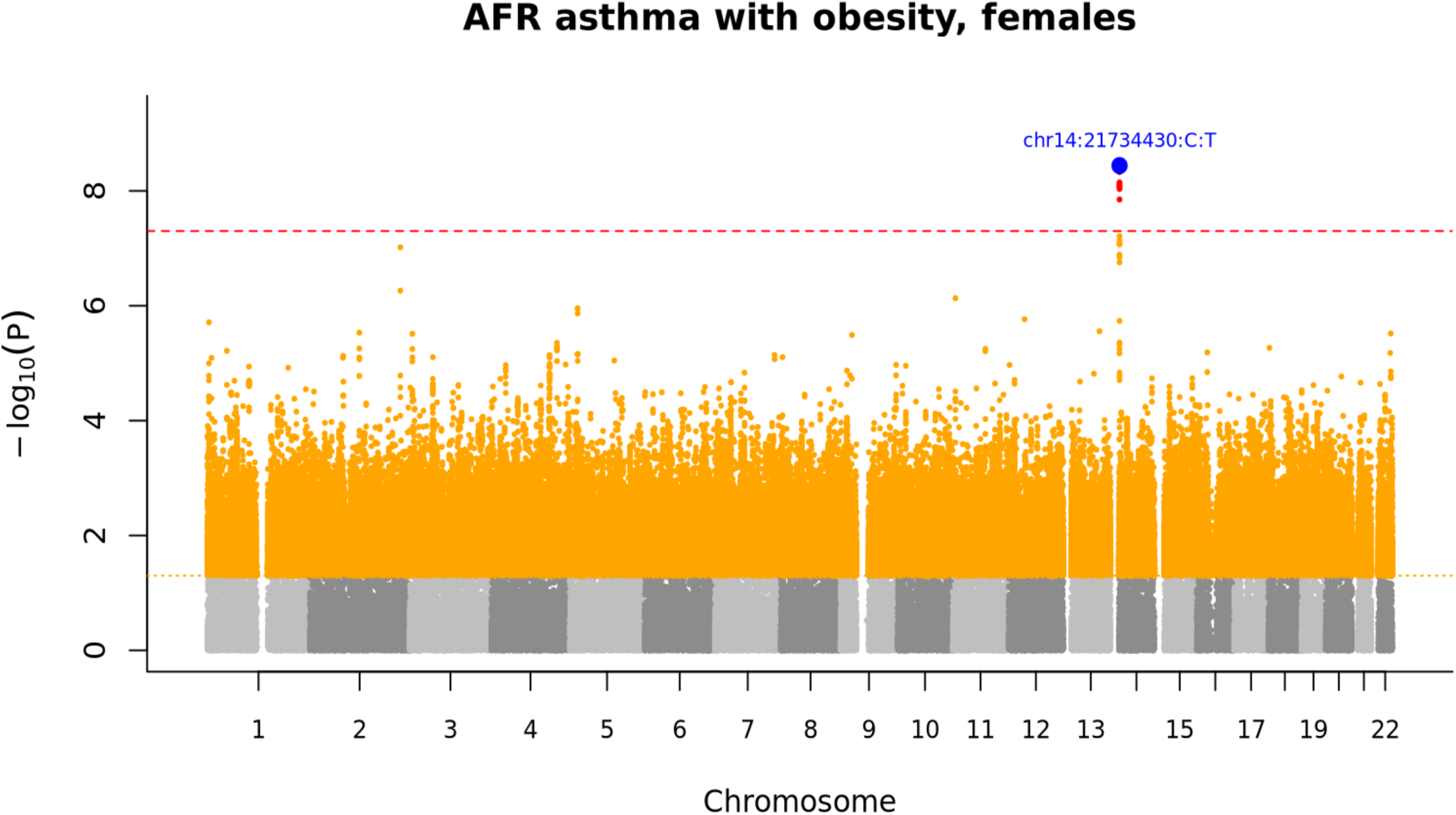
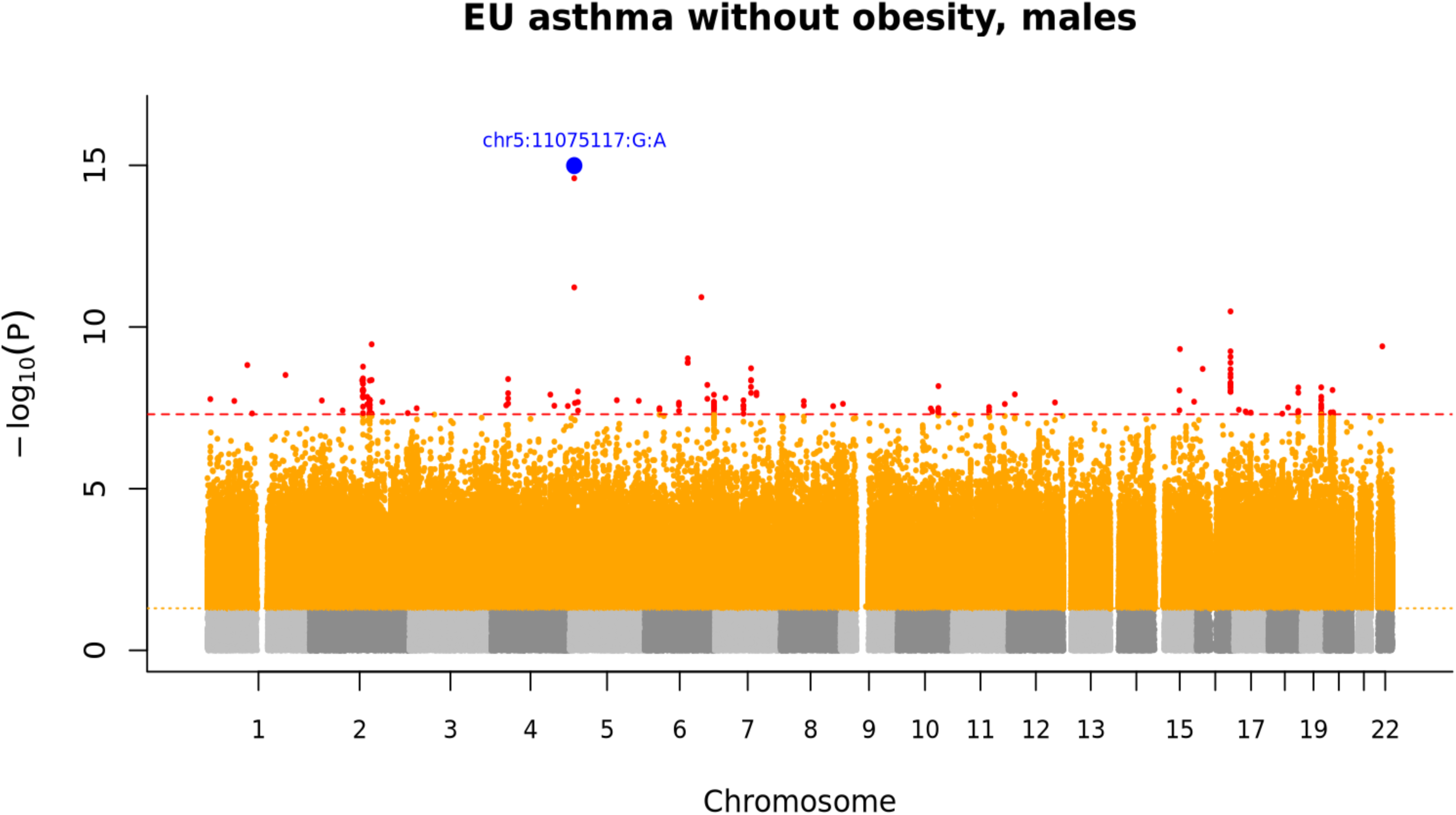
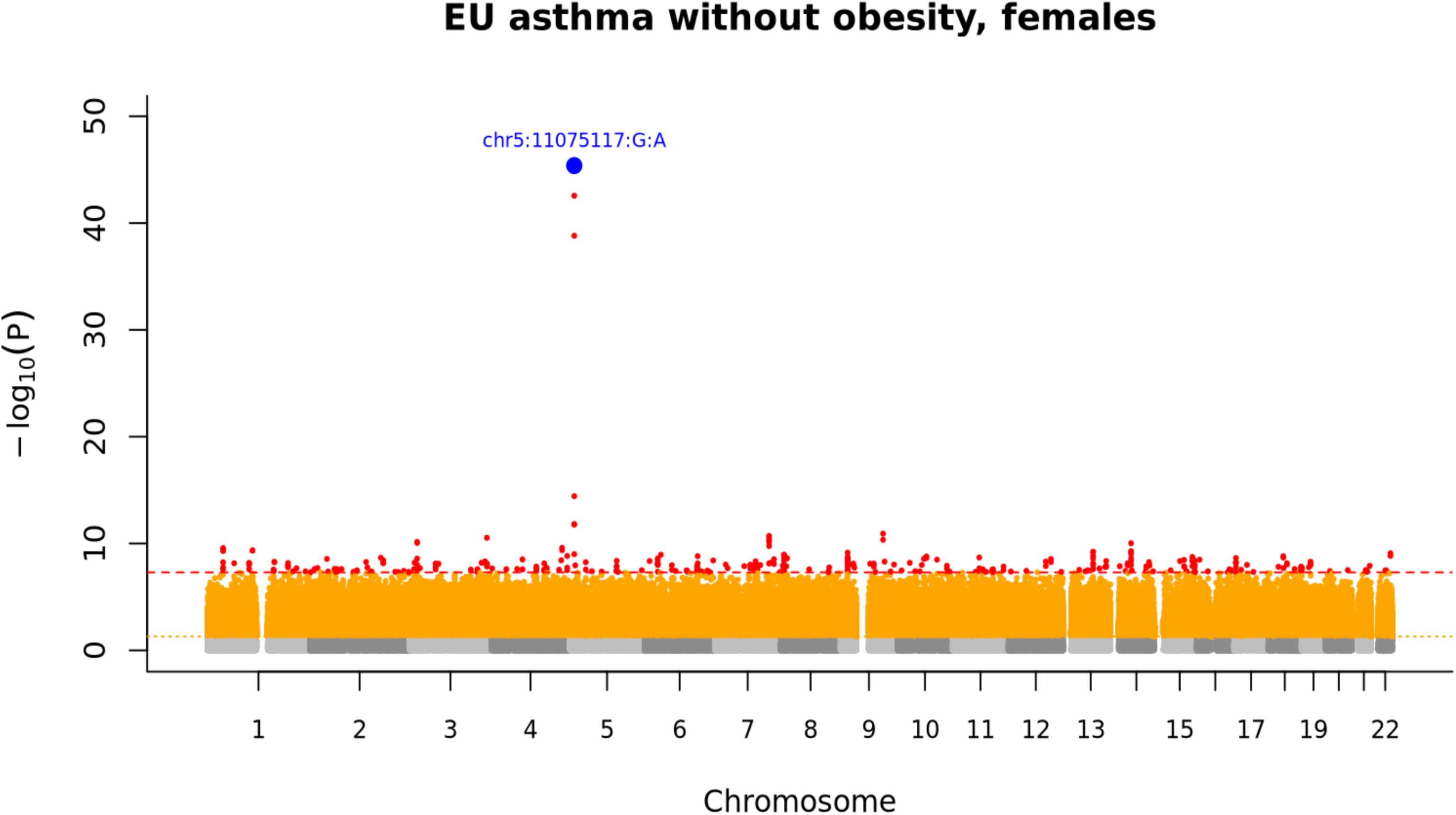
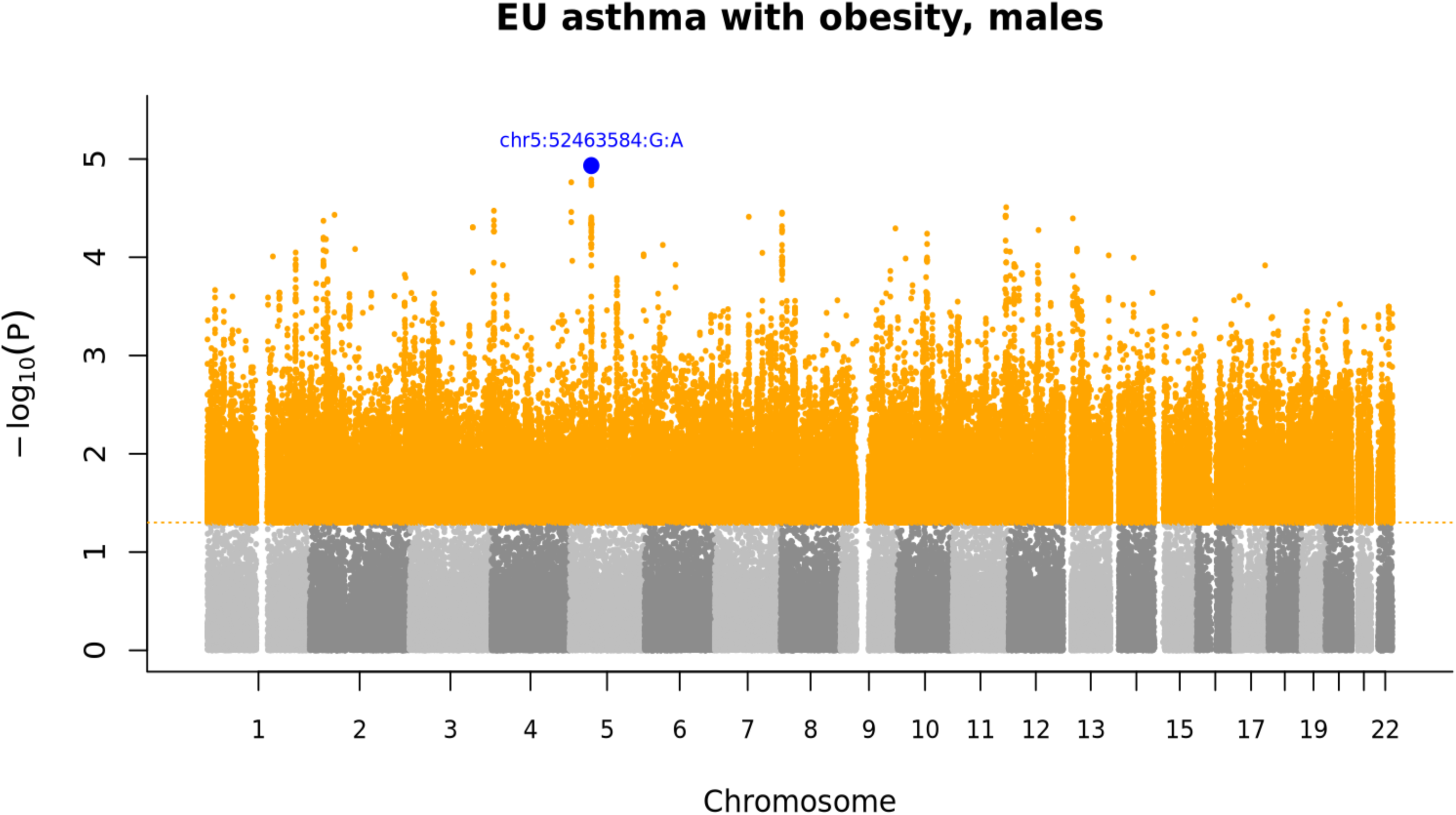
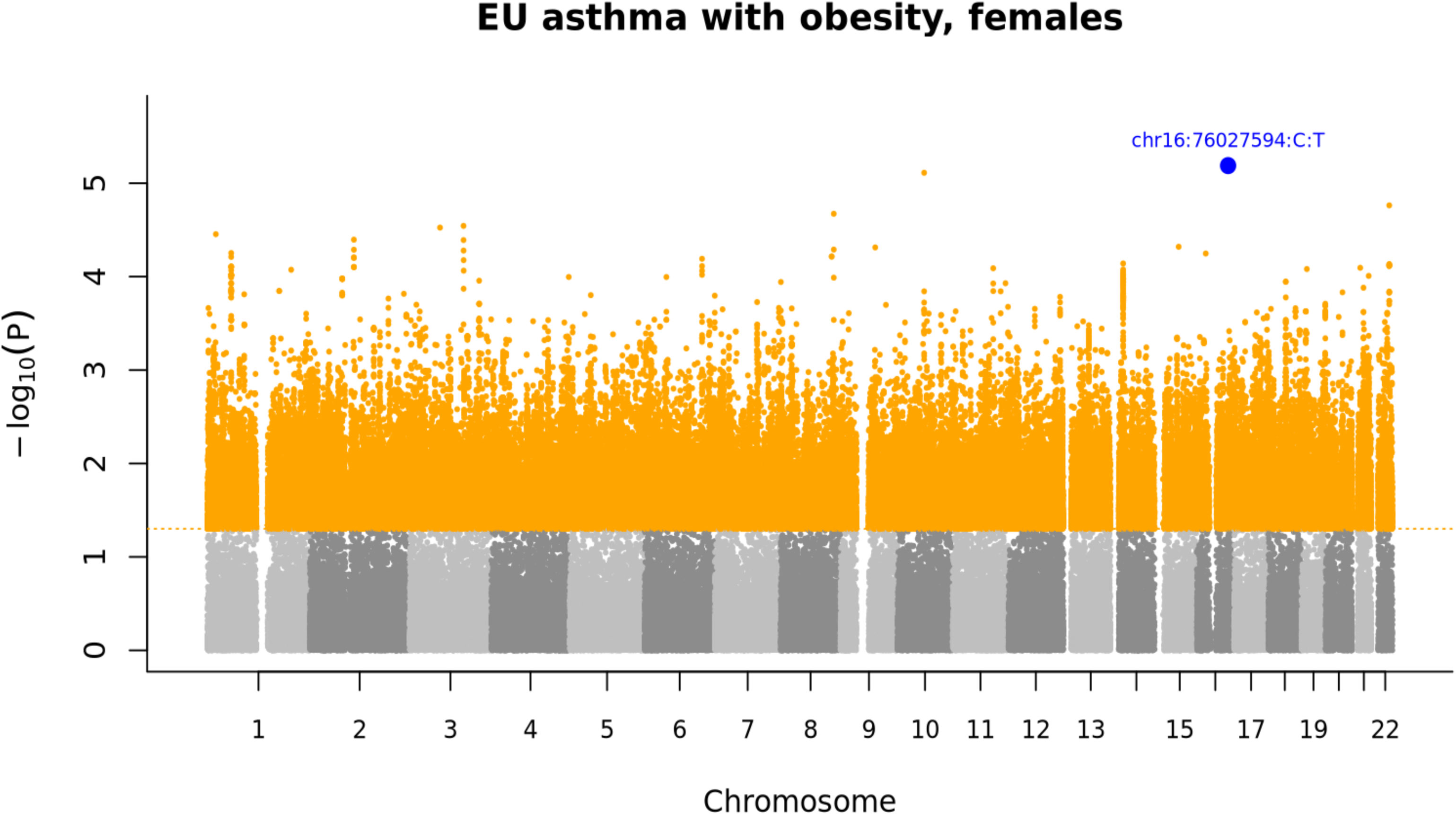
Manhattan plots for asthma GWAS meta-analyses. Manhattan plots show ancestry-specific GWAS meta-analysis results for asthma, asthma without obesity, and asthma with obesity in AFR and EU participants, including sex-stratified analyses for asthma without obesity and asthma with obesity. Each point represents a variant plotted by cumulative genomic position across chromosomes 1-22 on the x-axis and −log10(P) on the y-axis. Alternating grey colors indicate chromosomes, orange points indicate variants with P < 0.05, and red points indicate genome-wide significant variants with P < 5E-8. Dashed horizontal lines mark the genome-wide significance threshold (P = 5E-8) and nominal threshold (P = 0.05). The top variant in each plot is highlighted and labeled in blue.

**Table 1.** Asthma GWAS meta-analysis and lead loci.

| Ancestry / comparison | Variants analyzed | GWA significant variants | GWA 500-kb windows | Suggestive variants < 1E-6 | Suggestive 500-kb windows | Top signal | Shared loci |
| --- | --- | --- | --- | --- | --- | --- | --- |
| AFR | 18,063,038 | 7 | 2 | 159 | 63 | chr7:106018481:T:C; $\beta = 0.1168$, OR = 1.12, $P = 4.61E-10$ | 1 |
| EU | 9,995,105 | 147 | 25 | 595 | 162 | chr5:11075117:G:A; $\beta = 0.9952$, OR = 2.71, $P = 6.39E-59$ | 3 |

\* Genome-wide significant in one ancestry and nominal same-direction evidence in the other ancestry, $P < 0.05$ .

### Obesity-stratified asthma GWAS

Obesity-stratified analyses showed distinct association patterns for asthma with obesity and asthma without obesity (Table 2; Supplementary Table 5). Asthma with obesity showed no GWS variants in either ancestry. The strongest suggestive signal was chr2:219863806:C:T in AFR and chr11:12349420:T:C in EU. Asthma without obesity carried the GWS burden in both ancestries. In AFR, GWS lead loci included the strongest signal at chr6:153041377:T:C, followed by chr2:214337875:A:G and chr5:76396474:T:C. In EU, the strongest signal was chr5:11075117:G:A, followed by GWS lead loci on chromosomes 18 and 17.

**Table 2.**
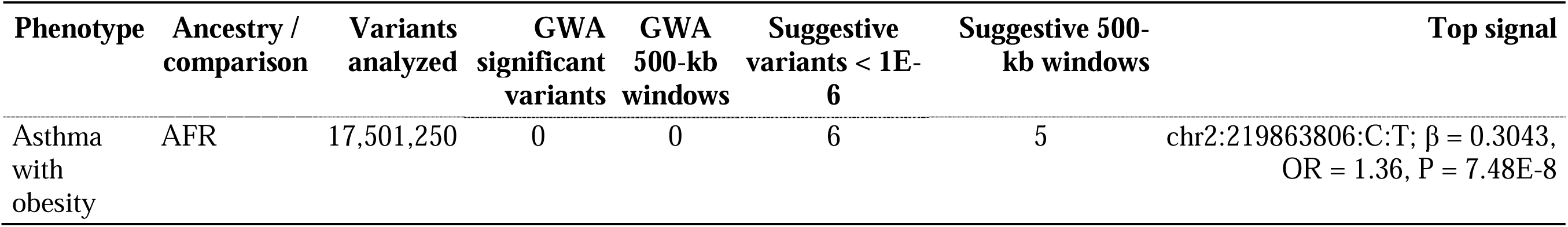
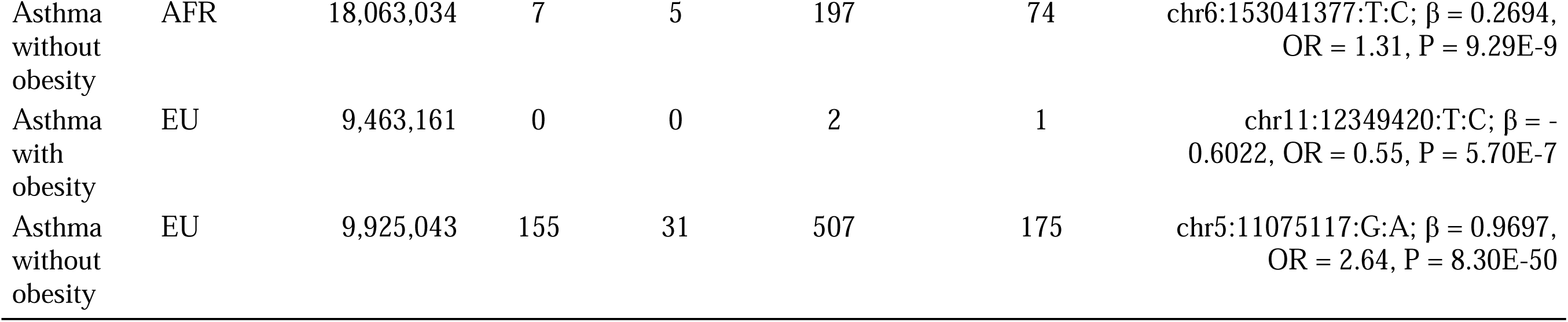
Obesity-stratified asthma GWAS meta-analysis and lead loci.

| Phenotype | Ancestry / comparison | Variants analyzed | GWA significant variants | GWA 500-kb windows | Suggestive variants < 1E-6 | Suggestive 500-kb windows | Top signal |
| --- | --- | --- | --- | --- | --- | --- | --- |
| Asthma with obesity | AFR | 17,501,250 | 0 | 0 | 6 | 5 | chr2:219863806:C:T; $\beta = 0.3043$, OR = 1.36, $P = 7.48E-8$ |
| Asthma without obesity | AFR | 18,063,034 | 7 | 5 | 197 | 74 | chr6:153041377:T:C; $\beta$ = 0.2694, OR = 1.31, P = 9.29E-9 |
| Asthma with obesity | EU | 9,463,161 | 0 | 0 | 2 | 1 | chr11:12349420:T:C; $\beta$ = -0.6022, OR = 0.55, P = 5.70E-7 |
| Asthma without obesity | EU | 9,925,043 | 155 | 31 | 507 | 175 | chr5:11075117:G:A; $\beta$ = 0.9697, OR = 2.64, P = 8.30E-50 |

Cross-ancestry sharing differed by obesity stratum. For asthma without obesity, two EU GWS lead variants showed nominal same-direction evidence in AFR: chr5:11075117:G:A and chr18:72820306:C:G. No AFR GWS lead variants for asthma without obesity showed nominal same-direction evidence in EU. Cross-stratum sharing was also observed, including an AFR asthma-without-obesity signal near *TSBP1-AS1*/*HLA-DRA* with nominal same-direction evidence in EU asthma with obesity, and multiple EU asthma-without-obesity signals at 17q21 with nominal same-direction evidence in AFR asthma with obesity and/or AFR asthma without obesity.

Obesity-stratified heterogeneity was evaluated within each ancestry by comparing asthma with obesity and asthma without obesity meta-analysis results. Among variants with significant obesity-stratum heterogeneity and GWS or suggestive evidence in at least one phenotype-specific analysis, 103 variants were retained in AFR and 22 in EU (Supplementary Table 6). In AFR, 5 were GWS for asthma without obesity; 5 variants were suggestive for asthma with obesity and 98 were suggestive or GWS for asthma without obesity. In EU, 4 were GWS for asthma without obesity; 2 variants were suggestive for asthma with obesity and 20 were suggestive or GWS for asthma without obesity. Most retained variants showed opposite directions of effect between asthma with obesity and asthma without obesity, including 102 of 103 variants in AFR and 21 of 22 in EU. No obesity-heterogeneous variants were shared between AFR and EU.

### Sex-stratified GWAS

Sex-stratified GWAS meta-analyses were conducted separately for asthma with obesity and asthma without obesity in AFR and EU participants (Table 3; Supplementary Table 7). In AFR asthma with obesity, no GWS variants were identified in males, although one variant reached the suggestive threshold of P < 1E-6; the strongest male signal was chr3:147092540:G:A (β = -0.367, OR = 0.69, P = 9.60E-7). In females, 9 variants reached GWS and 21 reached the suggestive threshold, corresponding to one GWS lead locus. The strongest female signal was chr14:21734430:C:T (β = 0.391, OR = 1.48, P = 3.63E-9).

**Table 3.** Sex-stratified asthma GWAS meta-analysis and lead loci.

| Phenotype | Ancestry / comparison | Sex | Variants analyzed | GWA significant variants | GWA 500-kb windows | Suggestive variants < 1E-6 | Suggestive 500-kb windows | Top signal |
| --- | --- | --- | --- | --- | --- | --- | --- | --- |
| Asthma with obesity | AFR | Male | 17,277,633 | 0 | 0 | 1 | 1 | chr3:147092540:G:A; $\beta$ = -0.3674, OR = 0.69, P = 9.60E-7 |
| Asthma with obesity | AFR | Female | 17,323,244 | 9 | 1 | 21 | 3 | chr14:21734430:C:T; $\beta$ = 0.3909, OR = 1.48, P = 3.63E-9 |
| Asthma without obesity | AFR | Male | 18,015,465 | 5 | 4 | 77 | 37 | chr16:89603181:G:A; $\beta$ = 0.5044, OR = 1.66, P = 3.07E-9 |
| Asthma without obesity | AFR | Female | 17,687,608 | 22 | 7 | 319 | 139 | chr2:241240094:C:T; $\beta$ = 0.5198, OR = 1.68, P = 1.45E-9 |
| Asthma with obesity | EU | Male | 9,385,500 | 0 | 0 | 0 | 0 | chr5:52463584:G:A; $\beta$ = 0.8007, OR = 2.23, P = 1.17E-5 |
| Asthma with obesity | EU | Female | 9,321,937 | 0 | 0 | 0 | 0 | chr16:76027594:C:T; $\beta$ = 1.4965, OR = 4.47, P = 6.48E-6 |
| Asthma without obesity | EU | Male | 9,692,231 | 169 | 61 | 1,109 | 344 | chr5:11075117:G:A; $\beta$ = 0.7520, OR = 2.12, P = 1.02E-15 |
| Asthma without obesity | EU | Female | 9,674,059 | 395 | 182 | 2,259 | 835 | chr5:11075117:G:A; $\beta$ = 1.3316, OR = 3.79, P = 4.15E-46 |

For AFR asthma without obesity, the male analysis identified 5 GWS variants and 77 suggestive variants, corresponding to 4 GWS lead loci. The strongest male signal was chr16:89603181:G:A (β = 0.504, OR = 1.66, P = 3.07E-9). The female analysis identified 22 GWS variants and 319 suggestive variants, corresponding to 7 GWS lead loci. The strongest female signal was chr2:241240094:C:T (β = 0.520, OR = 1.68, P = 1.45E-9).

In EU asthma with obesity, no GWS or suggestive variants were identified in either sex. The strongest EU male signal was chr5:52463584:G:A (β = 0.801, OR = 2.23, P = 1.17E-5), and the strongest EU female signal was chr16:76027594:C:T (β = 1.497, OR = 4.47, P = 6.48E-6).

For EU asthma without obesity, the male analysis identified 169 GWS variants and 1,109 suggestive variants, corresponding to 61 GWS lead loci. The strongest male signal was chr5:11075117:G:A (β = 0.752, OR = 2.12, P = 1.02E-15). The female analysis identified 395 GWS variants and 2,259 suggestive variants, corresponding to 182 GWS lead loci. The strongest female signal was also chr5:11075117:G:A (β = 1.332, OR = 3.79, P = 4.15E-46).

Sex heterogeneity was evaluated within each ancestry and obesity-stratified phenotype. Among variants with significant male-female heterogeneity and GWS or suggestive evidence in at least one sex-specific analysis, 19 variants were retained for AFR asthma with obesity, 331 for AFR asthma without obesity, none for EU asthma with obesity, and 2,106 for EU asthma without obesity (Supplementary Table 8). In AFR, retained heterogeneity signals were more common among females than males for both asthma with obesity and asthma without obesity. In EU, no retained sex-heterogeneous variants were observed for asthma with obesity. EU asthma without obesity showed 2,106 retained variants, including 90 male GWS variants and 327 female GWS variants. Most retained variants showed the same direction of effect in asthma without obesity, including 282 of 331 AFR variants and 1,910 of 2,106 EU variants.

GWS variants associated with more than one specific phenotype showed consistent directions of effect, primarily across EU asthma, EU asthma without obesity, and EU sex-stratified asthma without obesity analyses (Supplementary Table 9).

## Discussion

### Asthma genetic signals across obesity and sex strata

The overall asthma GWAS identified genome-wide significant loci in both AFR and EU ancestry populations, although the genetic architecture differed substantially between ancestries. AFR participants had two genome-wide significant loci, whereas EU participants had twenty-five, with only limited cross-ancestry replication. Specifically, one AFR locus and three EU loci showed nominal significance with concordant directions of effect in the alternate ancestry, highlighting both ancestry-specific and shared components of asthma susceptibility.

Stratification by obesity revealed an even more striking pattern. Genome-wide significant associations were almost exclusively observed among individuals with asthma without obesity, whereas no genome-wide significant loci were detected for asthma with obesity in either ancestry. Likewise, suggestive associations were consistently more numerous in asthma without obesity across both populations. These findings support the concept that obesity-associated asthma represents a biologically distinct phenotype in which environmental, metabolic, and inflammatory mechanisms may contribute more strongly to disease susceptibility than inherited common genetic risk. Obesity is known to influence asthma through multiple pathways, including altered respiratory mechanics, reduced lung volumes, systemic inflammation, adipokine dysregulation, and metabolic perturbations affecting airway biology (12). Together, our findings suggest that obesity substantially alters the genetic architecture of asthma and may attenuate the detectable effects of common susceptibility variants identified in non-obese asthma.

Sex stratification further refined the genetic architecture of obesity-defined asthma. GWS signals were identified in AFR females with asthma and obesity, whereas no GWS variants were detected in the corresponding combined-sex analysis, indicating female-specific genetic effects. In asthma without obesity, GWS signals were observed in both AFR and EU male and female strata. By contrast, no GWS variants were detected in AFR males with asthma and obesity or in either EU asthma-with-obesity sex stratum. The strongest asthma-associated variant in Europeans, chr5:11075117:G:A, remained the dominant signal in asthma without obesity and was the leading association in both male and female EU analyses, although its effect size was greater in females. Similarly, in AFR participants with asthma and obesity, sex-stratified analysis identified GWS locus in females but not in males, further supporting sex-specific genetic susceptibility.

Sex differences were most pronounced in asthma without obesity. Among AFR participants, females exhibited seven GWA windows compared to four in males, despite having fewer affected individuals. Likewise, in the EU cohort, females demonstrated 182 GWA windows compared with 61 in males, again despite fewer female cases. These findings suggest a substantially richer genetic architecture in females that is not explained by sample size alone. The observed sex-specific differences are biologically plausible.

Asthma prevalence shifts after puberty from predominance in boys to predominance in females and accumulating experimental and clinical evidence demonstrates that sex hormones influence both allergic and non-allergic airway inflammation, airway remodeling, and immune responses. These mechanisms may contribute to the enhanced genetic signals observed in females and highlight the importance of considering sex as a biological modifier in genetic studies of asthma (13).

### Shared and population-differentiated asthma association signals

#### Shared signals

Among variants shared between AFR and EU with the same direction of effect, the chr7 locus mapped to *CDHR3*. *CDHR3* is expressed in airway epithelium and functions as a receptor for rhinovirus C(14). Asthma-risk variation at *CDHR3* can increase rhinovirus C binding and replication in airway epithelial cells (15, 16). The same-direction association in AFR and EU is consistent with shared epithelial antiviral susceptibility.

The chr5 signal mapped to *CTNND2* showed the largest effect in both populations among the shared variants. *CTNND2* encodes δ-catenin, an adhesive-junction protein that participates in cadherin-catenin adhesion complexes and regulates dendritic spine and synapse morphogenesis during development(17). This signal links asthma to cell adhesion, cytoskeletal organization, and developmental/neural regulation.

The chr13 association signal lies near *FOXO1*, a transcription factor with immune relevance. Experimental studies have linked *FoxO1* activity to allergic airway inflammation, including macrophage polarization and type 2 inflammatory signaling (18).

The chr18 association signal annotated to *NETO1* had same-direction evidence in both populations. *NETO1* encodes a neuronal transmembrane protein involved in kainate-receptor function and synaptic transmission(19). This signal may point to neural regulation in asthma.

#### Heterogeneous signals

Among variants demonstrating ancestry-heterogeneous effects, the strongest association cluster localized to the 17q21 locus. Multiple variants within this region reached genome-wide significance in individuals of EU ancestry, whereas the corresponding associations were markedly attenuated or absent in AFR ancestry, indicating substantial ancestry-specific genetic effects. In particular, chr17:39851068:T:G in *IKZF3* showed opposite directions of effect between AFR and EU. This region includes *ZPBP2*, *GSDMB*, *ORMDL3*, and *IKZF3* and is one of the most reproducible asthma loci, especially for childhood-onset asthma (20). *ORMDL3* and *GSDMB* have been linked to airway inflammation and epithelial responses (20, 21). *IKZF3* encodes Aiolos, an Ikaros-family zinc-finger transcription factor that regulates lymphocyte development and B-cell differentiation(22). The weaker or opposite AFR association at 17q21 reflects differences in the asthma phenotype captured across populations, particularly if childhood-onset or virus-associated asthma contributed more strongly to the EU signal(23).

Heterogeneous association signals outside 17q21 involved additional immune and regulatory regions. A chr6 signal falls in the *HLA-DQA1*/*HLA-DQB1* region, where dense immune-gene structure and ancestry-specific LD can produce strong variant-level differences(24). The chr13 *FOXO1* and chr18 *NETO1* regions also appeared among cross-population signals.

#### Other loci

Association signals near *RORA* and *LY86*/*RREB1* point to immune regulation. *RORA* regulates type 2 inflammatory responses during pulmonary inflammation (25), while LY86 encodes MD-1, an RP105-associated regulator of TLR4 signaling and innate immune activation (26). *SATB1* links T-helper-cell differentiation and IL-5 regulation in type 2 immune programming (27). *ROBO2* and *PLXNA2* encode guidance-receptor pathways involved in axon guidance, cell migration, and cytoskeletal remodeling, which indicates airway structural and neurodevelopmental regulation (28). *MYCT1*/*SYNE1* points to endothelial-immune regulation and nucleo-cytoskeletal coupling. *MYCT1* regulates endothelial angiogenic behavior and immunity (29), while *SYNE1* connects the nuclear envelope to the cytoskeleton (30). *TENM2*, *APOL5*/*APOL6*, and *PLCXD3*/*OXCT1* point to synaptic organization, innate immune/lipid biology, and metabolic regulation (31, 32).

### Obesity-stratified asthma genetic signals

#### Shared signals

Shared association signals were identified only when ancestry and obesity status were crossed. An AFR asthma-without-obesity signal near *TSBP1-AS1*/*HLA-DRA* showed nominal same-direction evidence in EU asthma with obesity. *HLA-DRA* links this cross-stratum signal to CD4^+^ T-cell antigen recognition(33). Multiple EU asthma-without-obesity signals at 17q21 showed nominal same-direction evidence in AFR asthma with obesity and/or AFR asthma without obesity.

Within the same obesity stratum, sharing was limited to asthma without obesity. Two EU asthma-without-obesity GWS lead variants showed nominal same-direction evidence in AFR asthma without obesity, mapping near *CTNND2* and *NETO1* and involving epithelial-inflammatory susceptibility (17, 19).

#### Heterogeneous signals

Obesity-stratified heterogeneity was dominated by opposite directions of effect, indicating that obesity status changed allele behavior rather than only attenuating association. In AFR, heterogeneity association signals annotated to *RGS17*, *HLA-DRA*, *IQGAP2*, *SPAG16*, and *SV2C* involved airway signaling, antigen presentation, epithelial structure, ciliary function, and neural signaling, emphasizing airway response to the obese state. This is supported by the role of cAMP signaling in airway smooth-muscle function and by *IQGAP2*-mediated epithelial TLR4/NF-κB inflammatory signaling (34, 35). In EU, heterogeneity signals annotated to *APOE*, *CDH13*, *NETO1*, *NSUN7*, *SENP7*, and *CTNND2* involved lipid-related inflammation, adiponectin-related vascular or epithelial signaling, post-transcriptional regulation, and cell adhesion, emphasizing lipid-adiponectin and adhesion pathways at the metabolic-airway interface. *APOE* can regulate asthmatic inflammatory response (36), and *CDH13* encodes an adiponectin-binding protein that modifies allergic airway responses (37).

#### Other loci

In AFR without obesity, the association signals emphasized immune and airway-regulatory genes, including *RGS17*, *SPAG16*, *HLA-DRA*, *IQGAP2*, *MKLN1*, and *SV2C*. In EU without obesity, the signals were concentrated in canonical asthma regions, especially *ZPBP2*/*GSDMB*/*IKZF3*/*ORMDL3*, with additional signals at *NETO1*, *CTNND2*. These signals showed stronger alignment with established asthma loci, especially epithelial inflammation, antigen presentation, airway regulation, and immune-cell control.

### Sex-stratified asthma genetic signals

#### Shared signals

Shared male-female association signals were observed in asthma without obesity. In AFR asthma without obesity, shared signals included genes related to autophagy, vesicle trafficking, cytoskeletal organization, and lipid/RNA regulation, including *OPTN*, *SEPTIN2*, and *HDLBP*. *OPTN* links autophagy to NF-κB signaling (38), and septins organize cytoskeletal and vesicle-trafficking processes (39). In EU asthma without obesity, shared signals included genes linked to epithelial defense, extracellular matrix remodeling, neuronal growth-factor signaling, and cell migration, including *DMBT1*/*HTRA1*, *NRG3*, *HPSE2*, and *DOCK1*. *DMBT1* supports epithelial immune defense (40), *HTRA1* remodels extracellular matrix proteins (41), and *DOCK1* regulates cytoskeletal remodeling and migration (42).

#### Heterogeneous signals

Sex heterogeneity differed by obesity status and ancestry. In AFR asthma with obesity, the strongest heterogeneity association signals mapped near *OR4E1*/*LOC105370401* and were female-driven with opposite-direction effects in males. This points to a narrow female-specific obesity-related signal rather than a broad shared sex effect. In AFR asthma without obesity, same-direction female-stronger heterogeneity was enriched near *ATG10*, with additional signals near genes involved in cytoskeletal and membrane regulation. *ATG10* plays major roles in autophagy and stress-response(43).

In EU asthma without obesity, sex heterogeneity was broader and mostly same-direction with stronger female effects. Female-stronger signals were enriched near genes involved in neuronal synaptic signaling (*NPTX2*, *CTNND2*, *CSMD1*) (17, 44), epithelial integrity and extracellular matrix biology (*TNXB*) (45), microtubule organization (*CNTLN*) (46), and developmental regulation (*DACH1*, *CTNND2*) (17). Male-stronger signals included genes involved in extracellular matrix biology (SMOC2) (47), Wnt-related regulation (*DACT2*) (48), immune signaling and epithelial response (*PLCG2*) (49). Sex appeared to change the strength of airway, epithelial, immune, and neural asthma signals rather than creating completely separate male or female asthma pathways.

#### Other loci

Remaining sex-stratified GWA signals were concentrated in AFR female asthma with obesity and EU asthma without obesity. In EU asthma without obesity, female-specific signals again implicated neuronal and epithelial regulation near *NPTX2*/*TMEM130* (44), while male-specific signals included immune and regulatory genes such as *PLCG2*, *ABCB1*/*RUNDC3B*, *LIN28B*, and *DPYD*.

GWS variants associated with more than one phenotype showed consistent directions of effect, mainly across EU overall asthma, EU asthma without obesity, and EU sex-stratified asthma without obesity. This recurrence indicates that much of the stable EU asthma association signal was carried by asthma without obesity, especially at *CTNND2*, 17q21, and *NETO1*, reflecting baseline roles for cadherin-catenin adhesion/developmental regulation, epithelial-inflammatory susceptibility, and neural signaling.

## Conclusion

Asthma susceptibility is strongly modified by ancestry, obesity status, and sex, revealing distinct genetic architectures across clinically relevant subgroups. Shared signals near *CDHR3* and *FOXO1* across African and European populations support conserved roles for epithelial antiviral defence and immune regulation in asthma pathogenesis, whereas ancestry-heterogeneous associations at the 17q21 locus highlight population-specific genetic effects at a well-established epithelial-inflammatory susceptibility region.

Stratification by obesity and sex uncovered susceptibility loci that were masked in the combined asthma phenotype, demonstrating that these factors define the physiological and inflammatory context in which asthma risk alleles exert their effects. Asthma without obesity was enriched for genetic pathways involved in epithelial inflammation, antigen presentation, airway biology, and immune regulation, while sex-stratified analyses further identified developmental and neural signaling pathways, particularly among individuals without obesity. Together, these findings demonstrate that incorporating ancestry, obesity, and sex into genetic analyses substantially improves the resolution of asthma susceptibility loci and provides a framework for more biologically informed disease classification and precision medicine approaches.

## Acknowledgements

We thank all patients and their families who have participated in our research for the past two decades.

## Ethics approval and consent to participate

All experimental protocols were approved by the Institutional Review Board (IRB) of the Children’s Hospital of Philadelphia (CHOP) with the IRB number: IRB 16-013278. Informed consent was obtained from all subjects. If subjects are under 18, consent was also obtained from a parent and/or legal guardian with assent from the child if 7 years or older.

## Consent for publication

Not applicable.

## Data availability statement

GWAS summary statistics are provided in the Supplementary Tables. Full summary statistics will be deposited in the NHGRI-EBI GWAS Catalog. Additional information is available from the corresponding author upon reasonable request.

---

**Supplementary Table 1. AFR samples by genotyping/imputation batch and asthma phenotype**

**Supplementary Table 2. EU samples by genotyping/imputation batch and asthma phenotype**

**Supplementary Table 3. Asthma genome-wide significant and cross-ancestry loci**

**Supplementary Table 4. Asthma variants with significant AFR-EU heterogeneity and ancestry-specific genome-wide significant or suggestive associations**

**Supplementary Table 5. Obesity-stratified asthma genome-wide significant and cross-ancestry loci**

**Supplementary Table 6. Asthma variants with significant asthma-with-obesity versus asthma-without-obesity heterogeneity and phenotype-specific genome-wide significant or suggestive associations**

**Supplementary Table 7. Sex-stratified asthma GWAS lead loci by obesity status and ancestry**

**Supplementary Table 8. Asthma variants with significant sex heterogeneity and genome-wide significant or suggestive associations**

**Supplementary Table 9. Genome-wide significant variants associated with more than one specific asthma phenotype**

